# Scaling-Up the Impact of Teledermoscopy on the Early Detection of Skin Melanoma using Convolutional Neural Networks with Mobile Apps

**DOI:** 10.1101/2024.09.23.24314239

**Authors:** Tejasvi Tyagi, Swati Mathur Vempati, Kshitiz Upadhyay

## Abstract

Advances in the cloud technology for secured distributed data storage, modern techniques for machine learning (ML), and access to large populations through mobile apps provide a unique opportunity for the healthcare industry professionals in the areas of early screening and medical diagnostics for certain diseases. This research study demonstrates the potential of ML using convolutional neural networks (CNN) for medical diagnostics of skin melanoma. Specifically, a comparison is presented between a shallow CNN (3-layers) with Resnet50 (50-layers) to classify open datasets of skin melanoma images as malignant or benign. Various ML performance metrics such as accuracy, recall, precision and receiver operating characteristic (ROC) are presented to recommend a deep learning model for the mobile app. Also, a novel framework is proposed for the scalability and adoption of ML-based medical diagnostics by large masses as a mobile app running on data-secure cloud platform. Using the open datasets, it is shown that skin cancer can be accurately diagnosed with a mobile phone app while maintaining patient privacy and data security.

## 1. Introduction

The uncontrollable development of tissues in a specific body area is known as cancer. One of the most quickly spreading diseases in the world is skin cancer. Skin cancer is a disease in which abnormal skin cells develop out of control (**Fig. 1**). To determine potential cancer therapies, early detection and accurate diagnosis are essential. Melanoma, the deadliest form of skin cancer, is responsible for most skin cancer-related deaths in developed countries (www.cancer.org). It is caused when melanocytes, which produce pigment within skin, are hit with radiation. The melanocyte becomes cancerous and creates that tell-tale dark patch of skin. This lesion can either be malignant or benign. Malignant melanoma is constantly growing, and can progress through stages of cancer quickly, becoming very dangerous. Benign melanoma does not evolve and can be easily left alone. According to the Global Health Observatory (2022), Melanoma cancer is responsible for the most skin cancer-related deaths in the developed countries.

**Figure 1:**
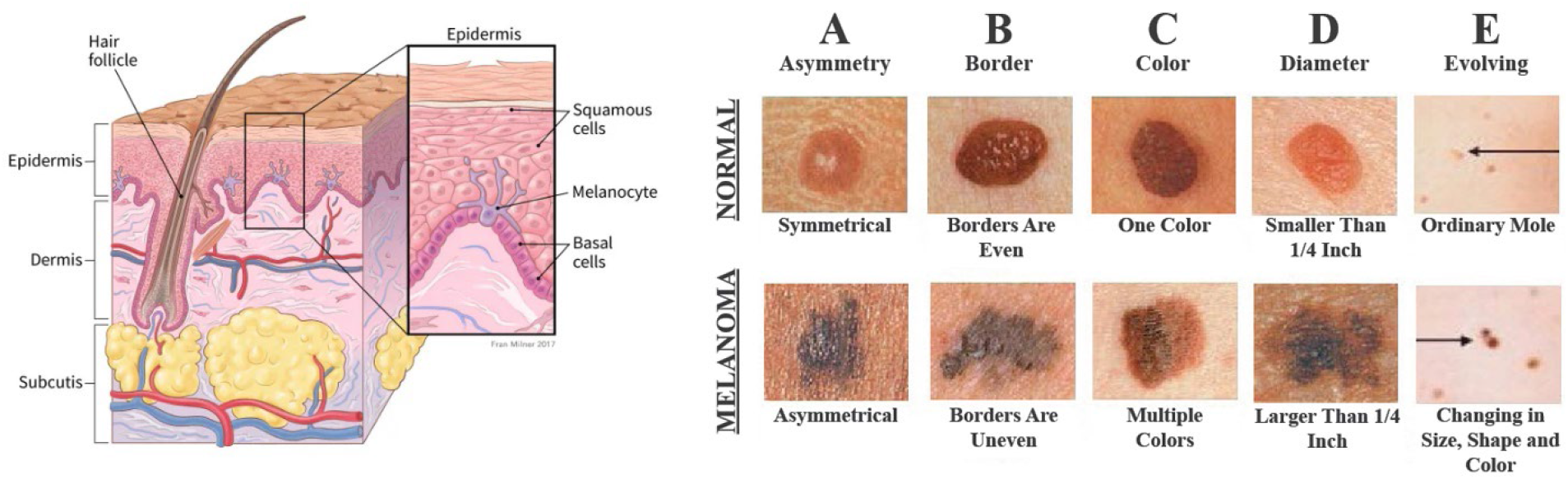
(Left) Labeled diagram of Skin Tissue (Skin Anatomy [2]); (Right) The A-B-C-D-E of diagnosing skin melanoma (Melanoma ABCDE [3]).

Artificial intelligence (AI) and machine learning (ML) have made significant progress by leaps and bounds in several areas such as self-driving or assisted-driving cars, computer vision, retail supply chain management, and healthcare. Computer ML models outperformed top human players in games like Chess and Alpha Go. A study by Weng et al. [1] has demonstrated that ML models have made significant improvements in the accuracy of cardiovascular risk prediction by increasing the number of patients identified who could benefit from preventive treatment while avoiding the unnecessary treatment of others.

Several prior studies [4-7] have investigated ML models for melanoma detection and a non-exhaustive review is presented here. Li et al. [8] advocated for construction of an artificial intelligence system in dermatology using skin image database through interdisciplinary collaborations. Bhatt et al. [9] presented a literature review of the state-of-the-art ML techniques (e.g. Support vector machine (SVM), K-Nearest Neighbors (KNN), and Convolutional neural networks (CNN) etc.) used to classify skin melanoma cancer as malignant or benign. Among open datasets reviewed, ISIC (2016) and ISIC (2018)/HAM10000 are also used in the present study.

Haenssle et al. [10] compared a Convolutional Neural Network (CNN) ML model’s diagnostic performance against a large international group of 58 dermatologists, including 30 experts, and reported that most dermatologists were outperformed by the CNN ML model. They recommended that irrespective of any physicians’ experience, they may benefit from the assistance by the image classification capabilities of a CNN. Reis et al. [11] developed InSiNet architecture that outperformed the other methods achieving an accuracy of 94.59%, 91.89%, and 90.54% in ISIC 2018, 2019, and 2020 datasets, respectively.

Delans et al. [12] presented “teledermoscopy” as an emerging technology for skin cancer detection and argued for teledermoscopy as an attractive tool to improve screening of skin cancers in populations with limited access to dermatological care. It may significantly reduce unnecessary biopsies through home monitoring of suspicious lesions and increase the overall cost-efficiency of dermatologic care by reducing the number of unnecessary in-person visits for clearly benign skin lesions. Wishma et al. [13] presented a methodology for building Android apps with associated services for teledermatology use cases of melanoma detection. However, their approach doesn’t address data security and patient privacy concerns.

The motivation of the present study takes inspiration from the above-mentioned impactful research works and aims to scale-up the usage of data-driven ML models in disease diagnostics through the contributions to datasets from large populations. To that end, a data-secure framework for mobile app is proposed that interfaces with a pre-trained ML model on image datasets to predict the skin melanoma classification. Due attention is paid to data security and patient privacy by using the advances in cloud technology to interface with mobile apps and running the ML model in a data secure environment. It is expected that early medical diagnostics or screening for skin melanoma can alleviate the burden on dermatologists in large populations with limited resources. Further, the data-driven ML approach continuously improves in the performance with increasing skin image datasets.

## 2. Materials and Methods

This research work uses data-driven ML techniques with mobile apps and hence, the main topics discussed are the deep learning framework, the statistical learning performance metrics, and mobile web app development. Description of the image datasets and CNN models is presented first. Then, several model performance metrics are defined. Lastly, a data-secure framework utilizing cloud technology with a mobile app is described.

### 2.1. Deep learning framework

Two CNN-based deep learning models are evaluated in this work for their performance of detecting melanoma: a shallow CNN and a pre-trained Resnet-50 model [14]. Both models are trained using about 8000 skin images classified as “benign” or “malignant” from the ISIC dataset and “Human Against Machine with 10000 training images” (HAM10000) dataset [15]. Overall, HAM10000 consists of 10015 dermatoscopic images of common pigmented skin lesions (**Fig. 2**).

**Figure 2:**
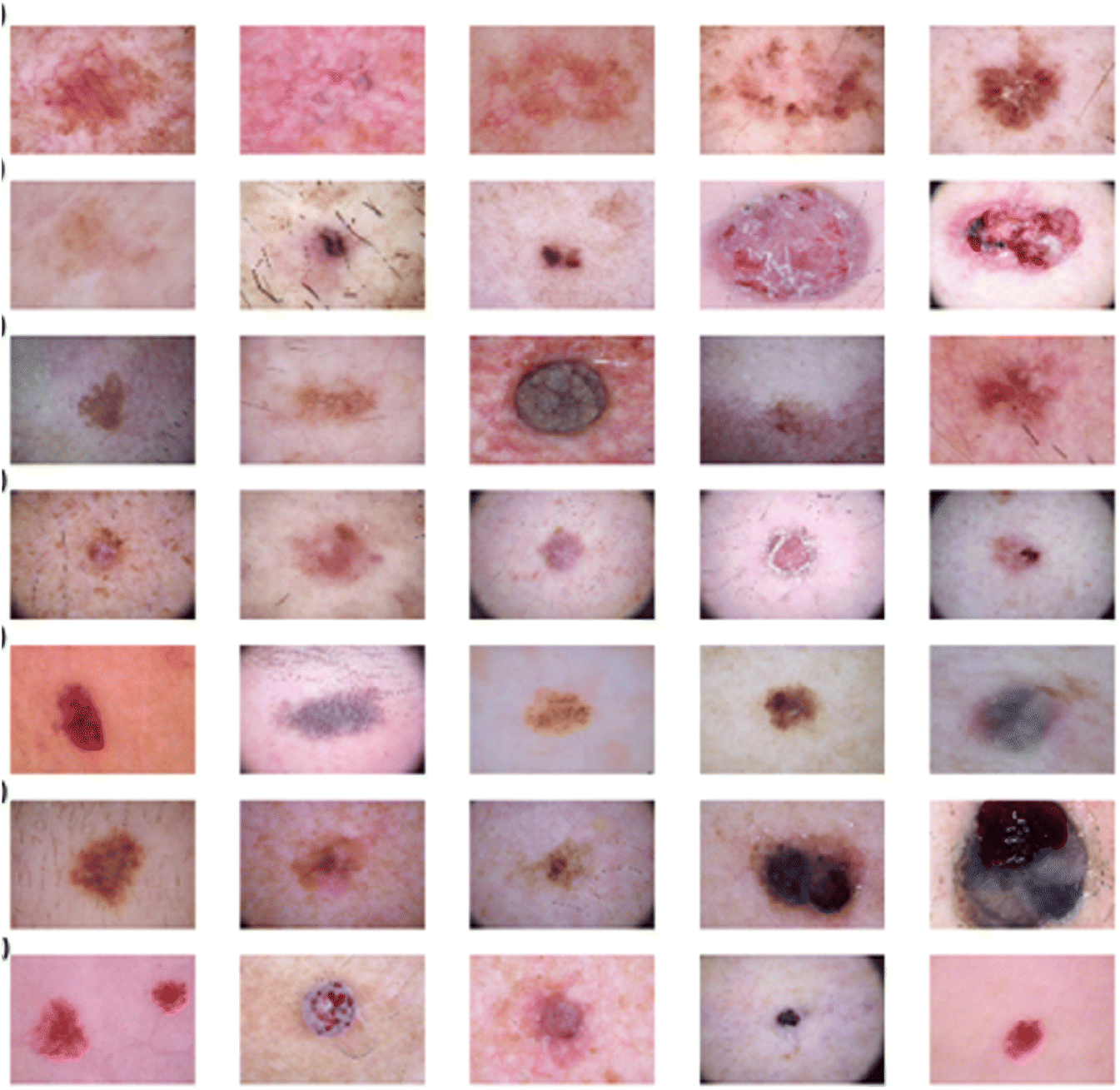
Example of images in the open dataset HAM10000 showing a wide range of skin cancers [15]. The present study deals with classification for only skin melanoma category.

A typical Convolutional Neural Network (CNN) architecture is shown in **Fig. 3 (Top)** with layers labeled as: Input, Convolution, Pooling, Flattening, Dense, and Output [16, 17]. The input layer represents the images as a combination of R, G, and B values at each pixel and thus, splitting an image into channels of three matrices with the corresponding values at the pixel location and the matrix dimensions corresponding to the image resolution. The role of activation functions in neural networks is like firing of neurons in the brain by transforming the weighted sum of all inputs into an output. Their non-linear behavior is essential for “learning” and typically, simple functions are used for easy training on large datasets. Examples include Tanh, Sigmoid, and ReLU. Convolution layers perform the tasks of feature extraction from a digitized representation of an image as a 2D array through a filter kernel and an activation function. Pooling operation performs down sampling operations on the feature maps obtained after the convolution layers that results in dimension reduction. Flattening layer transforms the output of last convolutional layer into a 1D array (i.e. vector) as an input layer for the fully connected neural network (dense layers) that is subsequently trained using the backpropagation algorithm. Dense or fully connected layers exploit the backpropagation algorithm for training the weights and biases in the network for the “reduced” dataset in the form of features instead of images. Lastly, the output (Softmax) layer is used for the label model prediction output [18-22].

**Figure 3:**
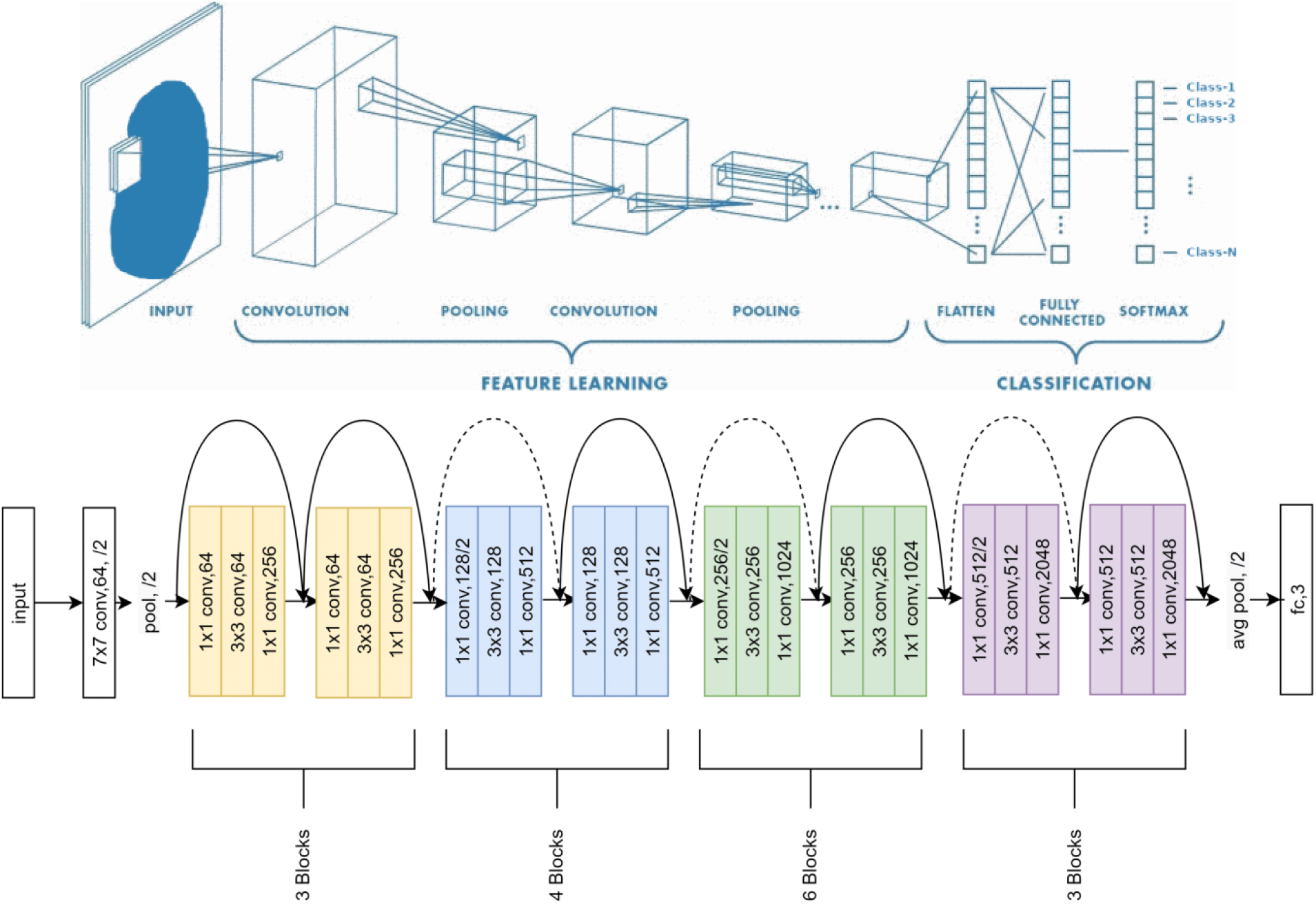
(Top) Workflow of a convolution neural network (CNN) showing various layers with their roles for the classification tasks on image datasets (Adapted from Vocaturo et al. [23]); (Bottom) the ResNet 50 Architecture [14, 24].

In this work, a shallow CNN is used first to learn about image datasets and computational resources. Images were reduced to 224 × 224 × 3 arrays. 2D convolutional layer, followed by max pooling, and dropout layers were stacked twice. It is followed by flattening, dense, and output layers. In our study, the two-state output will be text labels as “Malignant” or “Benign”. Other ML model parameters were: ReLU for activation functions, Adam optimizer in backpropagation, epoch of 50, and batch size of 64.

ResNet is a deep CNN architecture introduced by He et al. [24] and builds on four repetitive components – Convolution layers, Convolution blocks, Residual blocks, and Fully connected layers. ResNet-50 is a specific implementation that uses 3-layer convolution blocks of different sizes in the sequence of 3-, 4-, 6-, and 3 blocks (**Fig. 3, Bottom**). The novel idea of using residual blocks with “skipping” connections allow the network architectures to be deep and yet mitigate the problem of vanishing gradients during the model training [14, 24]. In this study, a pre-trained ResNet model in Keras was used with skin image datasets. Other ML model parameters for ResNet were: ReLU for activation functions, Adam optimizer in backpropagation, epoch of 50, and batch size of 64.

### 2.2. Statistical analysis

Overfitting is a critical concern when deep learning models are used on limited datasets. It occurs when a model aligns too closely with the training dataset and exhibits a large increase inerrors when making predictions on data other than that used in the training process. Ripley [25] describes the data split into training-validation-testing sets as: “Training set — A set of examples used for learning, that is to fit the parameters of the classifier, Validation set — A set of examples used to tune the parameters of a classifier, for example to choose the number of hidden units in a neural network, and Test set — A set of examples used only to assess the performance of a fully-specified classifier”. In this study, testing datasets were never used for tuning hyperparameters and therefore, provided an unbiased estimate of the generalization error. In this work, we avoid overfitting by dividing the dataset into separate training and testing sets (80:20 data splits) and choosing the number of epochs (=50) that minimize testing loss. Further, a cross-validation approach was also used for shallow CNN model with 3 folds in image dataset to confirm that similar level of ML model predictive accuracy was achieved.

Several performance metrics are used to evaluate the two trained CNN models. A confusion matrix is a tabular representation of the actual data against model prediction for classification tasks (**Fig. 4**).

**Figure 4:**
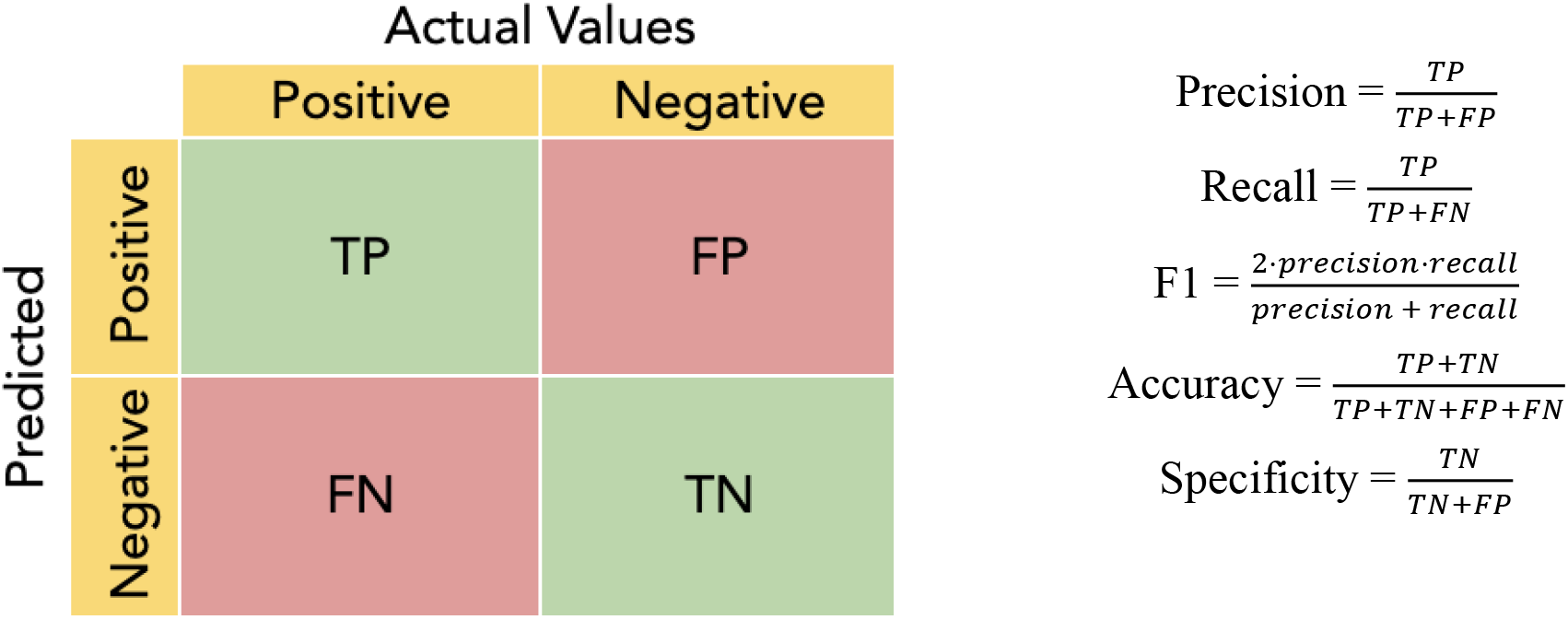
(Left) A confusion matrix for classification problem; (Right) Different ML model performance metrics.

Specifically, the following definitions are used in defining the confusion matrix based on True Positives (TP), True Negatives (TN), False Positives (FP), and False Negatives (FN): Accuracy is the fraction of true predictions (both positive and negatives) out of all the data. Precision is the fraction of true positives out of all positives predicted by the model. Recall is the fraction of model predicted positives out of total positives and is the same as sensitivity or true positive rate (TPR). F1 is the harmonic mean of precision and recall. Specificity is the fraction of model predicted negatives out of the total negatives. Receiver Operating Characteristic (ROC) is the curve obtained by plotting TPR (recall) against FPR (1 – specificity) at different classification thresholds and helps determine which model learns better.

### 2.3. Mobile web application development

Cazzaniga et al. [26] presented a teledermatology system cycle and recommended the use of ML-based apps to make it cost-effective (**Fig. 5**). Medhat et al. [27] presented a comparative study for skin melanoma detection using MobileNet-V2 among three different CNNs on mobile phones.

**Figure 5:**
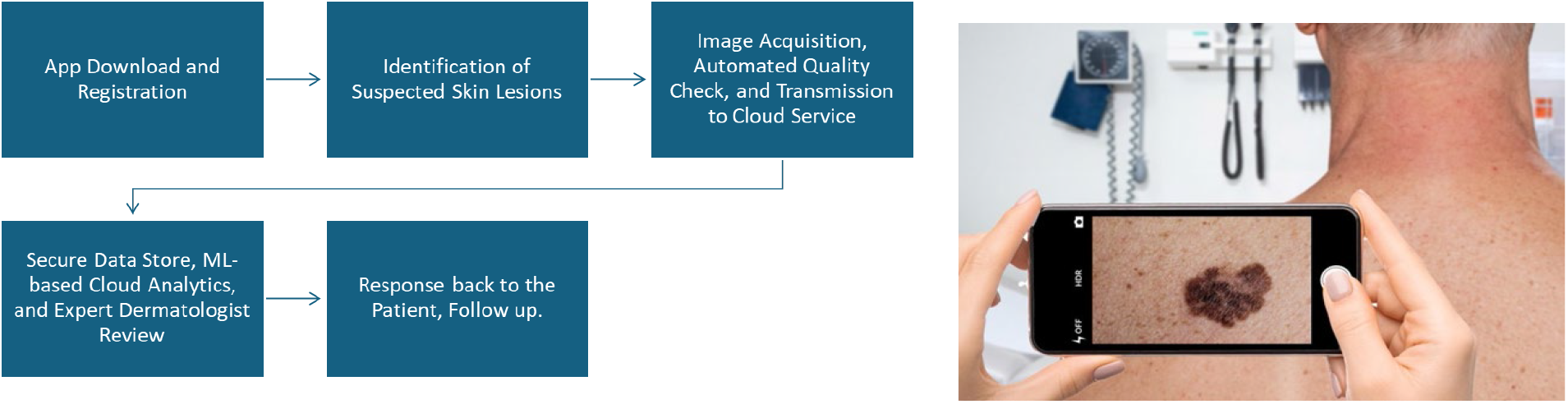
(Left) Teledermatology System Cycle adapted from Cazzaniga et al. [26]; (Right) Connecting mobile apps to machine learning models for the early detection of skin cancer can help scale-up the service for large populations. [28]

A typical electronic health record (EHR) management system using cloud server addresses the patient privacy required for HIPPA compliance and data security required by the hospitals and healthcare providers (**Fig. 6**). The user password provides the required privacy for the patient-specific information to be uploaded to the data-secure cloud server and the output of the ML model is then sent back to the patient. Liu et al. [29] advocated the use of cryptography (such as Blockchain) to further enhance data security for large biomedical databases.

**Figure 6:**
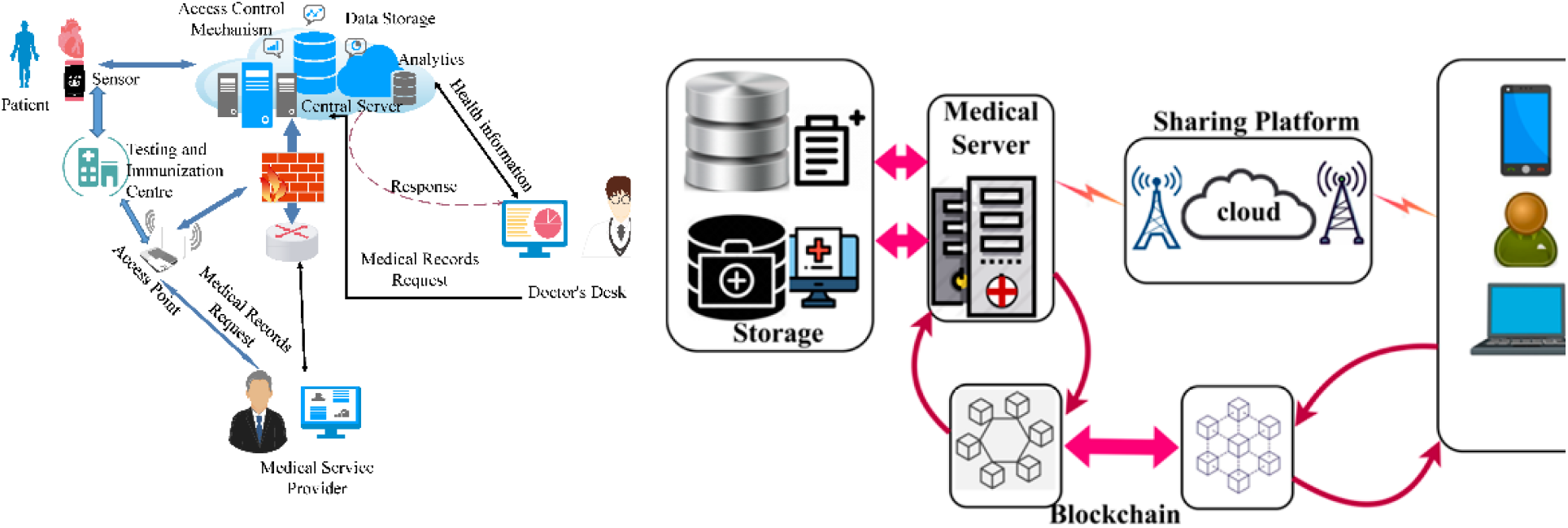
(Left) Cloud-centric EHR management system architecture [29]; (Right) Proposed scheme for improving biomedical data security with blockchain [28].

## 3. Results and Discussion

The confusion matrix and receptor-operator curve from the CNN model are shown here to illustrate the evaluation of model performance metrics (**Fig. 7**). It is observed that a shallow CNN model achieved the prediction accuracy of 83.7% accuracy. The ROC curve shows that this model learned quickly and can be useful for early diagnostics. However, the false negatives (Recall or F1) must be reduced further since their consequences can be severe for the patients.

**Figure 7:**
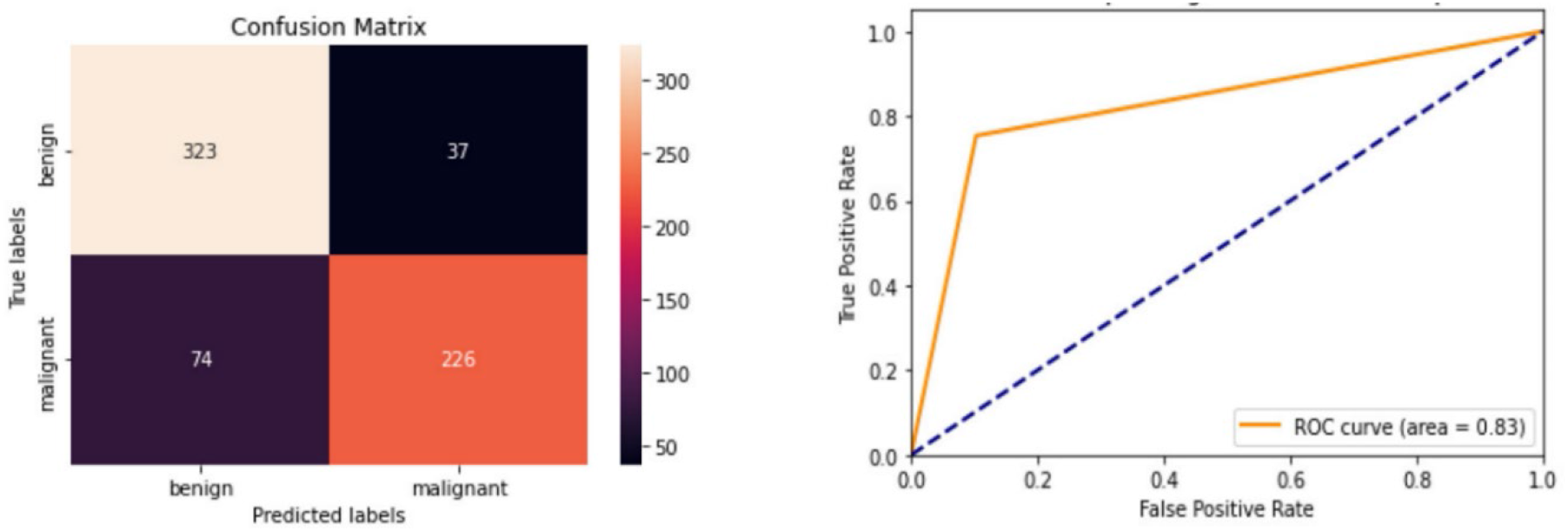
(Left) Confusion matrix for the CNN model on the test dataset; (Right) Receiver Operating Characteristic (ROC) for the CNN model.

For comparison, two CNN models with different architectures (due to differences in number of layers, activation functions, and hyperparameters during training) are studied further. The shallow CNN model (3 convolution layers) reached around 65% accuracy on average, while the Resnet50 model had 96% accuracy during training and over 80% in testing datasets (**Fig. 8, left**). Loss function values decrease with increase in epochs during training for both models. However, the loss function values increase with epochs for test datasets with Resnet50 (**Fig. 8, right**).

**Figure 8:**
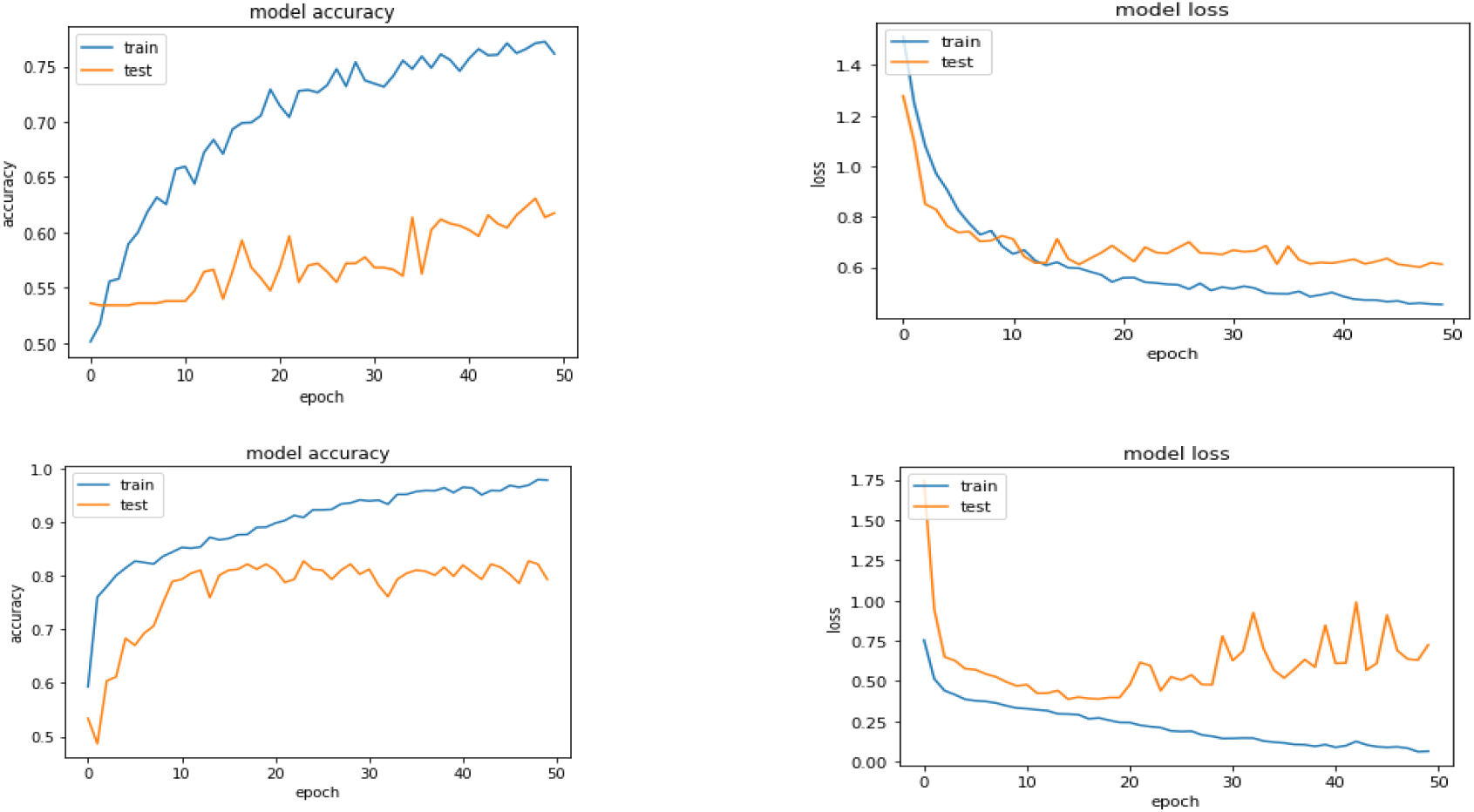
Improving model performance metrics using (Top row) a simple CNN and (Bottom row) ResNet50: (Left) Model accuracy vs. epochs; (Right) Training and testing loss function values vs. epochs.

Lastly, a framework is proposed for the adoption of ML models for medical diagnostics using a mobile app on the cloud platform. We implemented the code as a mobile/web application and ported it to the cloud platform for ease of accessibility while maintaining patient privacy and data security. The HIPAA-compliant mobile app is a patient portal that allows users to capture and maintain their skin images and submit them. The images submitted are securely uploaded to the cloud server where the image is processed based on the AI algorithms. The patient will be provided with updates on their submitted skin images, or they will be asked to contact the provider. A history of the images submitted is maintained to keep track of them in the app. The patient can maintain their profile and other settings in the app (**Fig. 9**).

**Figure 9:**
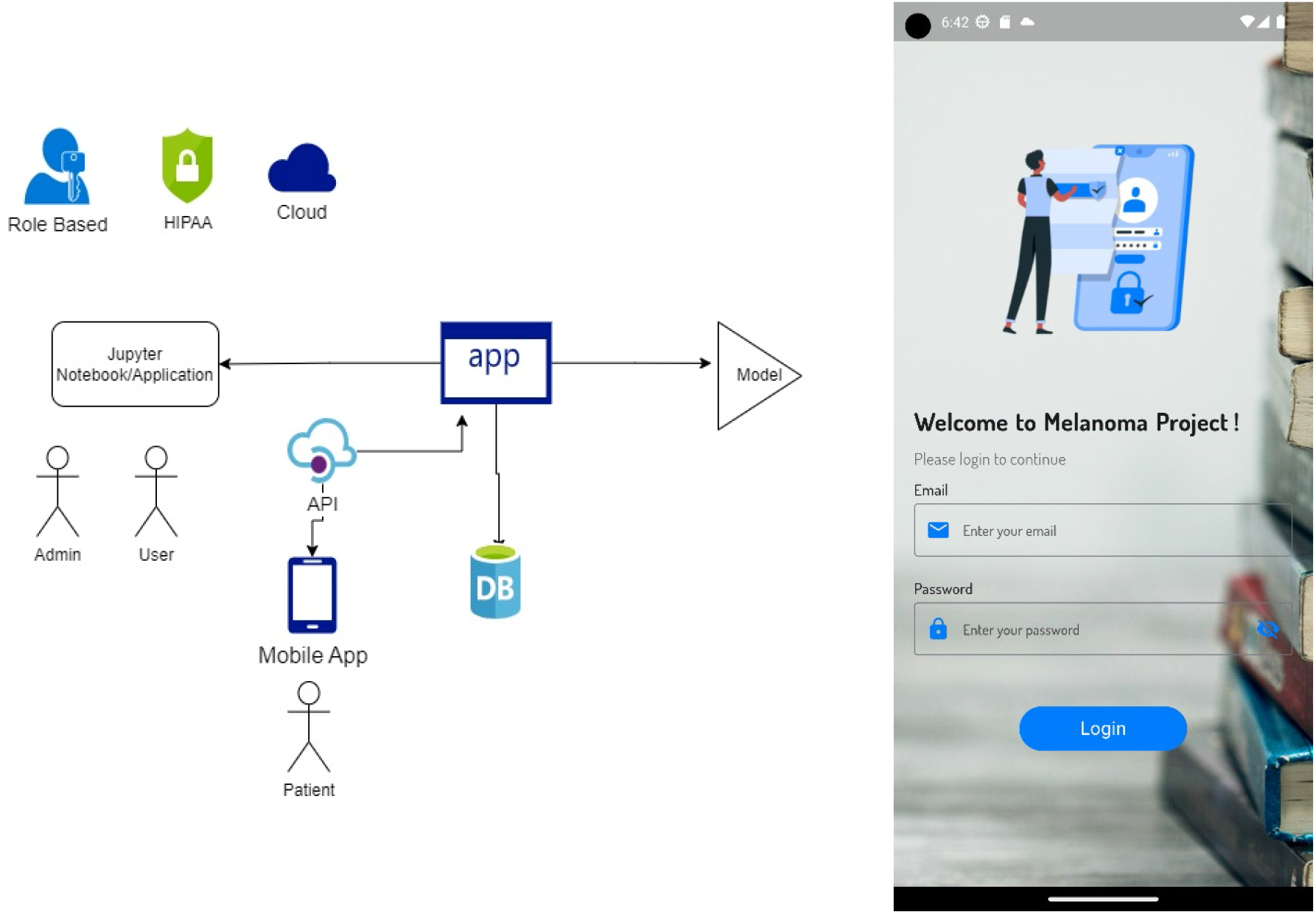
(Left) Proposed architecture for the data-secure mobile app; (Right) Screenshot of login page of the developed mobile app for the cloud-based security in the present work.

## Conclusion and Future Work

The present study used open datasets and code repositories to compare ML models of varying levels of fidelity. Specifically, image datasets for skin melanoma were used to classify the image as benign or malignant. A deep CNN (ResNet50) was shown to improve training and test prediction accuracies and F1-scores for classification of skin melanoma.

At present, the adoption of ML models for medical diagnostics is hindered by the concerns of patient privacy and data security. To that end, a framework is proposed using the cloud platform along with a mobile app. It is argued to be a scalable approach because mobile phones have already penetrated a large portion of the population around the world.

## Data Availability

All data produced in the present study are available upon reasonable request to the authors.

## Acknowledgments

First author (T.T.) would like to thank Prof. M. Tyagi (LSU) for his advice and supervision on data science and machine learning topics, and Mr. Deepak Borole (Chetu, Inc.) for the help in designing a mobile app that is HIPPA compliant. Readily searchable stock images are used from a wide range of websites for compiling the information and explanation of various technical terms.

## Disclaimer

An earlier version of this research project was presented at the 5th Annual Meeting of the SIAM Texas-Louisiana Section, November 4-6, 2022 (Tyagi [31]). All the datasets and associated code/app are developed using open-source tools [32] and will be made available to the readers upon request to the authors and posted on the repository maintained by the first author (T.T.). The original data source is available at https://dataverse.harvard.edu/dataset.xhtml?persistentId=doi:10.7910/DVN/DBW86T.

The related Data paper can be found at doi:10.1038/sdata.2018.161.

